# A biologically annotated neural network for proteomic discovery in Parkinson’s disease

**DOI:** 10.64898/2026.04.29.26351681

**Authors:** Avish Vijayaraghavan, Lorin Crawford, Krishnakant Saboo, Ava P. Amini, Ashley Mae Conard, Abby L. Olsen, Lana M. Chahine, Kristen A. Severson

**Affiliations:** Microsoft Research, Cambridge, MA USA; Imperial College London, London, UK; University of California San Francisco, San Francisco, CA USA; Department of Neurology, University of Pittsburgh, Pittsburgh, PA USA

## Abstract

Machine learning models that can utilize high-dimensional data to make predictions and derive biological insights can improve understanding of diseases. Here, we develop a biologically annotated neural network model for proteomics data (P-BANN) which has several practical advantages: (1) it incorporates known relationships between proteins and signaling pathways into its architecture design; (2) it uses Bayesian principles to enable variable selection on the most important proteins for a disease of interests; and (3) it combines structured and black-box variational inference to analyze different classes of phenotypes at scale. To demonstrate the value of the approach, we apply P-BANN to one of the most common neurodegenerative disorders: Parkinson’s disease (PD). We consider two biomarker-defined phenotypes within the PD population: presence of neuronal-predominate aggregated *α*-synuclein in cerebrospinal fluid, and changes in dopamine transporter binding in the striatum on imaging. By considering biomarkers of both neuropathological hallmarks of PD, we can examine the extent to which their underlying biology is connected. Using the P-BANN framework, we discover sparse, statistically-calibrated sets of proteins which map to pathways, enabling more straightforward interpretation and generation of testable hypotheses.

## 1 Introduction

As the availability of high-throughput proteomic testing expands, so too does the need for techniques that can utilize these high-dimensional data to make predictions and derive insights to improve disease understanding and intervention. Several works have previously demonstrated the value of proteomic studies in uncovering novel biology [35, 21, 25]. However, these studies must deploy a series of disjointed analyses to uncover these insights. This ad-hoc landscape can be replaced with a single technique so long as it fulfills certain criteria. Specifically, the single approach should be able to incorporate prior biological knowledge, be applicable to small datasets, and perform protein subset discovery with well-calibrated evidence. To meet these needs, we propose a novel approach, which we refer to as a proteomic biologically annotated neural network, or P-BANN.

In many machine learning applications, large datasets allow black-box models to discover complex relationships. However, in biomedical research, disease prevalence may be low and/or data collection costly, making such datasets unattainable. Our approach offsets these limitations by leveraging biological priors to enhance learning from small samples. One manner in which past work in genomics applications has proposed to integrate biological information is via model architecture design [32, 63, 20, 14, 17]. In these approaches, instead of using dense, fully connected networks, sparse connections based on genomic relationships (e.g., gene networks or signaling pathways), are used. For instance, Elmarakeby et al. [17] propose an approach, called pathway-aware multi-layered hierarchical network (P-NET), using pathway annotations from the Reactome [18] database to define neural network connections. However, utilizing these annotations alone may not be sufficient to yield interpretable results as the prediction may depend on a complex and non-linear combination of all input measures. Often the goal of high-throughput -omics assays is hypothesis generation (i.e., the selection of a small number of candidates for measurement or intervention, sometimes called subset identification in the machine learning literature). As a workaround to enable subset identification, Elmarakeby et al. [17] use a post-hoc explainable model called DeepLIFT [47] to calculate importance scores to rank input genes based on their influence on model output. Explainability models depend on post-hoc justifications that often lack statistical calibration, such as effect sizes tied to a null hypothesis, and may not faithfully recover the true decision boundary of a black-box predictor. In contrast, interpretable models are structured with intrinsic statistical parameters, like sparsity or biologically inspired modularity, enabling calibrated hypothesis testing, verified effect sizes, and transparent feature-to-output mappings [12]. Towards that end, in Demetci et al [14], the authors propose an inherently sparse model that depends on only a small number of inputs. They introduce biologically annotated neural networks (BANNs) where the hidden layer of the neural network is designed to capture single nucleotide polymorphism (SNP)-set effects using the NCBI’s Reference Sequence database [41] combined with Bayesian priors which lead to interpretable significance measures.

In our work, we seek to extend BANNs to a class of flexibly specified phenotypes, i.e. remove the requirement that the outcome variable is binary. We chose the BANNs model as the starting point for several reasons. First, it has already been demonstrated to capture biological priors. Second, the authors consider non-linear interactions, which are likely important for protein effects. And finally, they provide well-calibrated posterior inclusion probabilities (PIPs), a measure of how likely a model weight is non-zero and contributes to the model output, a key statistic for subset learning from high-dimensional data. Our novel model differs from the previous BANNs model in several ways. First, we use pathway-annotations as the source of biological prior. Second, we use horseshoe priors instead of spike-and-slab, which is more suited to the expectation of a larger range of effect sizes of proteins compared to SNPs. Third, we propose the use of black-box variational inference, specifically for the calculation of the intractable observed-data log-likelihood. This change enables flexible swapping of phenotype distributions without the need to do detailed derivations.

To demonstrate the value of the P-BANN model for protein subset identification and experimental hypothesis generation, we focus on applications in Parkinson’s disease (PD). PD is one of the most common neurodegenerative disorders and carries high morbidity, as well as substantial financial and societal costs [60]. The PD population is highly heterogeneous in presentation and progression and spans a complex mix of sporadic, rare, and common gene variant associations [31, 5]. The prevalence of PD is increasing, but therapies that modify the disease course are lacking [49]. PD is typically diagnosed via clinical criteria, but the recent availability of *in vivo* biomarkers has enabled a move toward a biological definition and characterization of the disease. Notably, cerebrospinal fluid (CSF) *α*-synuclein seed amplification assay (CSF*α*synSAA) [48, 37] and dopamine transporter (DAT) single-photon emission computed tomography (SPECT) binding imaging [36] are thought to capture the core pathophysiology of Parkinson’s disease: pathogenic aggregation of *α*-synuclein (*α*syn) protein and loss of dopaminergic neurons in the susbtantia nigra [15].

The majority of individuals with a diagnosis of PD based on clinical criteria also meet the biomarker-based definitions. However, there are individuals who clinically exhibit typical levodopa-responsive parkinsonism and demonstrate evidence of dopaminergic dysfunction on imaging, but who do not demonstrate abnormal CSF*α*synSAA. About 7% of individuals with sporadic PD belong to the latter group [48], but this is particularly common among individuals with certain genetic forms of PD. For example, over 35% of individuals with *LRRK2*-associated parkinsonism are negative for CSF*α*synSAA (CSF*α*synSAA-) [39, 28, 46, 48]. Whether these cases do not have abnormal *α*syn, or rather have forms that are not detected by current *in vivo* assays or immunohistochemical methods remains to be fully clarified.

Clinically, people with *LRRK2*-associated parkinsonism who are CSF*α*synSAA-are largely indistinguishable from individuals who are positive (CSF*α*synSAA+), though as a group they show some differences [8, 28]. The few cases that have undergone post-mortem examination demonstrate evidence of other neurodegenerative pathologies, namely Alzheimer’s disease (AD) type tau (3R and 4R), and less commonly hyperphosphorylated tau or TAR DNA-binding protein 43 (TDP43) [1, 10, 46]. Whether there are molecular differences that can be detected *in vivo* in *LRRK2*-associated parkinsonism cases who are CSF*α*synSAA- and CSF*α*synSAA+, beyond measures of aggregated *α*syn, remains unknown. In addition, the extent to which such molecular differences may correlate with other PD biomarkers is unknown. Addressing these key knowledge gaps will increase our understanding of the pathophysiology of PD, inform more precise biologic definitions of the disease, and aid in targeted therapeutic development.

While PD is common among older adults, it is rare at the population level, and genetic forms of PD constitute 10-15% or less of PD cases [59]. Thus, any dataset consisting of *LRRK2* parkinsonism cases is expected to be of small sample size, especially for studies that have acquired biomarker data such as CSF and imaging. This combination of inherently small cohorts, combined with complex biology and opportunities for better patient stratification and therapeutic discovery is an ideal setting to demonstrate the utility of the P-BANN model.

We use a dataset of proteomic measures to investigate the relationship between protein expression and biomarkers for a cohort of *LRRK2*-associated parkinsonsim. Our approach enables us to discover pathway-based subsets of proteins which predict *in vivo* biomarkers. Because of the ability to flexibly define phenotype, our approach can be applied both to the binary CSF*α*synSAA and temporal DAT SPECT measures. Our contributions are as follows. First, we develop the P-BANN model described above. Then, we apply the model to the cohort, considering separately CSF*α*synSAA status and DAT SPECT trajectory. Finally, we analyze and contextualize the results in the current literature.

## Results

### 2.1 Developing a biologically annotated neural network for proteomics

The protein biologically annotated neural network (P-BANN) model is designed to discover a sparse set of proteins and protein interactions that are predictive of a particular phenotype (see Fig. 1). This is achieved via a Bayesian neural network where the model architecture is defined based on biological pathway data. The priors over the weights are selected such that many values are likely to be zero. Briefly, we created a model of phenotype in which the distributional parameters are defined by a BANN, which in turn depends on protein expression (see eqn. 1). Biological pathway data defines a sparse set of connections from the inputs (protein expression) to the hidden layer (pathways). The hidden layer nodes are then connected to the phenotype (outcome). Horseshoe priors were selected for all weights as they are the continuous sparsity-inducing prior and can be written using Gaussian scale mixtures, enabling structured variational inference (see Methods Sec. 4.2 for further detail). The posterior parameter estimates are learned by optimizing the evidence lower bound, employing black-box estimates for the data log-likelihood term (see Methods Sec. 4.3). These choices enable flexible adaptation to many distributional choices of phenotype. After training, the posterior inclusion probabilities (PIPs; see Eqn. 10) are used to interpret the model. The PIPs provide calibrated estimates of the likelihood that a weight is non-zero. By incorporating the sparse structure in the architecture design, the PIPs of the weights that connect the hidden layer to phenotype can be contextualized based on the pathway they represent.

**Figure 1.**
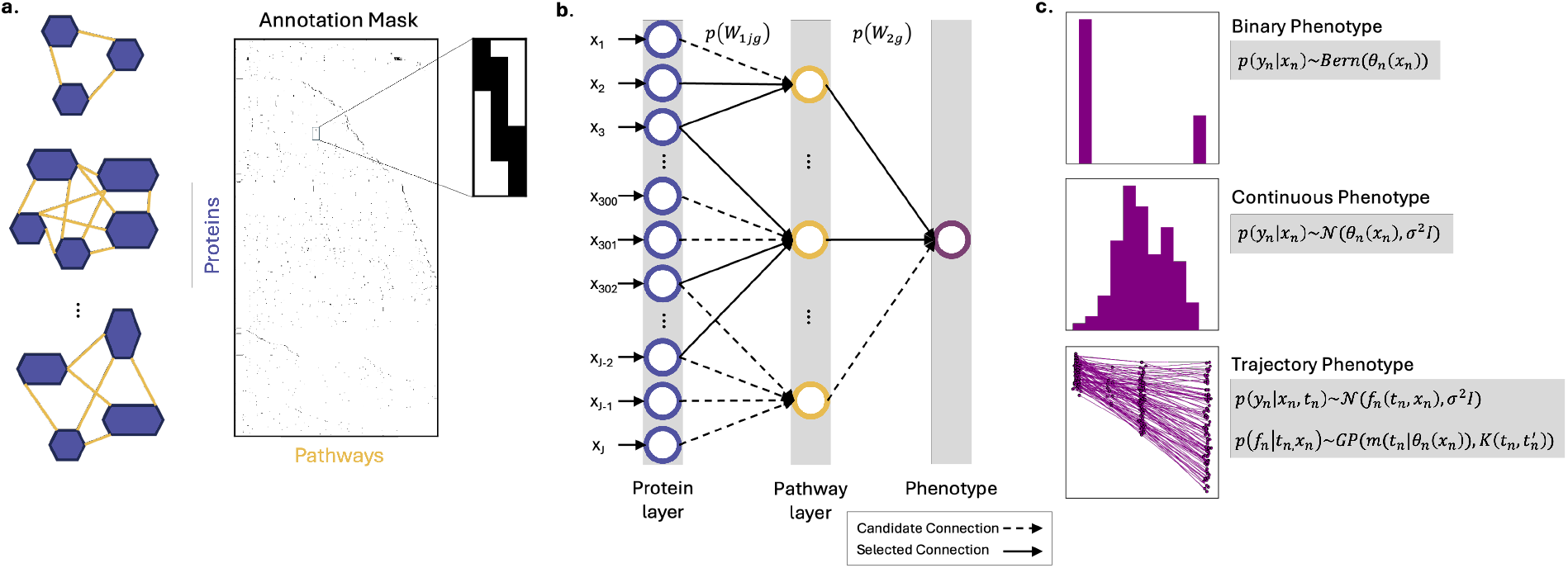
Overview of the protein biologically annotated neural network approach.**a.**Biological annotations, such as pathway definitions from the Reactome database, are used to create an annotation mask, capturing protein-protein interactions. **b**. The annotation mask is used to define the connections in a sparse Bayesian neural network. Sparsity-inducing priors are used to enable subset discovery. **c**. Phenotype can represent a variety of candidate distributions such as binary, continuous, and trajectory due to the use of black-box variational inference.

### 2.2 P-BANN discovers proteomic signatures in *LRRK2* biomarkers

To apply P-BANN to PD, we selected a sample of *LRRK2*-associated-parkinsonism patients from the Parkinson’s Progression Markers Initiative (PPMI) cohort [33] (demographics: Table 4, details: Sec. 4.1). As noted in the introduction, CSF*α*synSAA status and DAT SPECT binding imaging are thought to capture the core pathophysiology of PD. Participants in the PPMI cohort that we included in our analysis have each of these measures along with proteomics data, as measured by SomaScan obtained at their baseline study. Using these data, we define two related P-BANN models each corresponding to one of the aforementioned biomarkers. We use annotations from the Reactome [18] database to map the measured proteins to pathways (see Fig. 1a). This establishes the sparse connections from the inputs to the hidden layers (see Fig. 1b). The same annotations, which we refer to as the annotation mask, are used for both settings.

The first setting uses CSF*α*synSAA status (specifically positive or negative) as the outcome. This phenotype is modeled using a Bernoulli distribution and includes age and sex as possible confounders. The data is divided in training and testing splits. The CSF*α*synSAA P-BANN achieved 81% and 82% accuracy on the train and test splits, respectively. This predictive performance corresponded to 8 unique pathways and 15 proteins. Among the identified pathways, 5 pathways had a PIP greater than 0.6, indicating a strong predictive probability. 2 pathways had a PIP between 0.1-0.6, indicating intermediate predictive probability. The fifth pathway had a PIP < 0.1. The complete PIPs are reported in Table 1. PIPs ≤ 10^−3^ are treated as zero.

**Table 1.**
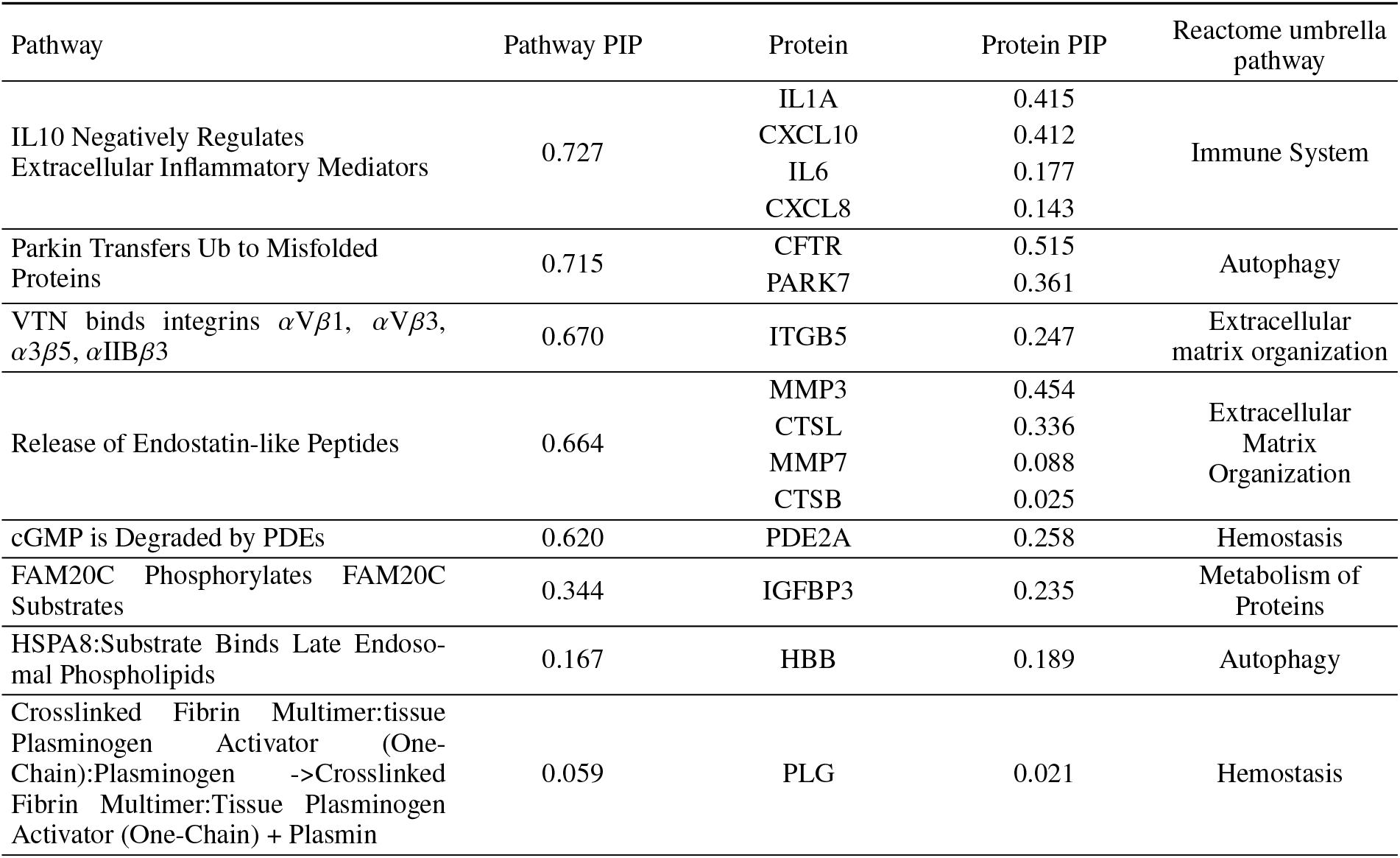
Pathway and protein posterior inclusion probabilities (PIPs) for the CSF*α*synSAA status prediction task.

The second setting used the mean striatum trajectory from DAT SPECT. The trajectory is modeled using an amortized Gaussian process, where the P-BANN predicts the hyperparameters of the prior mean function. Given the absence of strong evidence for higher order effects, we selected a linear mean as a function of time. The intercept and slope of the mean function are each modeled with P-BANNs. Again using a train and test split, without consideration for CSF*α*synSAA status, the model achieves a root mean square error of 0.24 and 0.25, respectively. The model identified seven unique pathways and eleven proteins that were associated with baseline mean striatum intensity. Two pathways had a PIP greater than 0.6, and the remaining five had a PIP between 0.1-0.6 (Table 2). We then identified 24 unique pathways and 36 proteins that were associated with changes in mean striatum intensity over time (Table 3). 9 pathways had a PIP > 0.6, 12 had a PIP between 0.1-0.6, and 3 had a PIP < 0.01.

**Table 2.** Pathway and protein posterior inclusion probabilities (PIPs) for the mean striatum at baseline prediction task.

| Pathway | Pathway PIP | Protein | Protein PIP | Reactome umbrella pathway |
| --- | --- | --- | --- | --- |
| pro-HD5 is Cleaved by Trypsin | 0.732 | PRSS2 | 0.092 | Immune System |
| Recruitment of Rnd1 to Plexin A | 0.612 | RAC1 | 0.35 | Developmental Biology |
| NOTCH1 Binds DLL/JAG Ligand in Cis | 0.397 | NOTCH1 | 0.353 | Signal Transduction |
| VTN Binds Integrins αVβ1, αVβ3, α3β5, αIIBβ3 | 0.351 | ITGB5 | 0.294 | Extracellular Matrix Organization |
|  |  | ITGB1 | 0.121 |  |
|  |  | ITGB3 | 0.063 |  |
| Mast Cell Carboxypeptidase hydrolyzes Angioten... | 0.319 | AGT | 0.579 | Metabolism of Proteins |
| HSPA8 Binds Substrate | 0.227 | PARK7<br>HBB<br>PLIN3 | 0.297<br>0.08<br>0.07 | Autophagy |
| Phosphorylation and Activation of PLCG | 0.148 | FCGR2A | 0.254 | Immune System |

**Table 3.** Pathway and protein posterior inclusion probabilities (PIPs) for the mean striatum trajectory BANN.

| Pathway | Pathway PIP | Protein | Protein PIP | Reactome umbrella pathway |
| --- | --- | --- | --- | --- |
| FGFR3b Binds to FGF | 0.89 | FGF20<br>FGF9<br>FGFR3 | 0.094<br>0.044<br>0.027 | Signal Transduction |
| Active FLT3 binds to CSK | 0.847 | FLT3 | 0.067 | Immune System |
| IFN $\alpha/\beta$ binds to IFNAR2 | 0.821 | IFNB1<br>IFNA2<br>IFNA4<br>IFNA8 | 0.269<br>0.196<br>0.182<br>0.033 | Immune System |
| IL21:IL21R:JAK1 binds IL2RG:JAK3 | 0.786 | IL21R<br>IL2RG | 0.037<br>0.031 | Immune System |
| HSPG2 (Perlecan) Degradation by MMP3, Plasmin, (MMP12) | 0.768 | PLG | 0.018 | Extracellular matrix Organization |
| COL1A2 Gene Expression is Stimulated by p-2S-SMAD3:p-2S-SMAD3:SMAD4 | 0.763 | SMAD3 | 0.018 | Gene Expression/Signal Transduction |
| GPVI binds Fyn and Lyn | 0.696 | FYN | 0.137 | Hemostasis |
| PRDM4 (SC1) Binds to p75NTR | 0.665 | PRDM4 | 0.158 | Signal Transduction |
| PI3K Binds ICOS | 0.659 | ICOSLG | 0.119 | Immune System |
| ppDVL Recruits RAC | 0.595 | DVL2<br>RAC1 | 0.058<br>0.024 | Signal Transduction |
| IL12B Binds IL23A | 0.582 | IL23A | 0.322 | Immune System |
| NPM1-ALK-dependent MAPK8,9 phosphorylation | 0.547 | MAPK9<br>NPM1 | 0.121<br>0.021 | Disease |
| ICAM1-5 Bind Integrin αLβ2 (LFA-1) | 0.534 | ICAM2<br>ICAM1 | 0.192<br>0.059 | ECM/Immune System |
| SIRT1 Deacetylates HSF1 | 0.527 | HSPA1A | 0.158 | Cellular Response to Stimuli |
| IL15 Binds IL2RB:JAK1 and IL2RG:JAK3 | 0.317 | IL2RB | 0.235 | Immune System |
| CETP-Mediated Lipid Exchange: LDL Gains Cholesterol Ester | 0.23 | APOF | 0.014 | Transport of Small Molecules |
| ARL2:GTP Bind PDE6D on KRAS4B | 0.226 | ARL2 | 0.358 | Signal Transduction |
| Cathepsin G hydrolyzes Angiotensin-(1-10) to Angiotensin-(1-8) | 0.219 | AGT<br>GZMH<br>CTSG | 0.374<br>0.266<br>0.231 | Metabolism of Proteins |
| FRS2 Binds to Active TrkA Receptor | 0.121 | FRS2 | 0.017 | Signal Transduction |
| RUNX2 Gene Expression from Proximal (P2) Promotor is Stimulated by ESR1:estrogen, ESRRA:PPARG1CA, TWIST1 and RUNX2-P2, and Inhibited by ESRRA:PPARG1CB and DEXA:NR3C1 | 0.118 | PPARGC1A | 0.121 | Gene Expression (Transcription) |
| Interaction of Nectins with Afad | 0.115 | NECTIN1 | 0.56 | Cell-cell Communication |
| LPL Hydrolyses TGs from Mature CMs | 0.091 | GPIHBP1 | 0.334 | Metabolism/Sensory Perception |
| GHBP Binds GH | 0.028 | GH1<br>GHR | 0.094<br>0.031 | Immune System |
| SARS-CoV-1 nsp1 Binds to PPIase | 0.02 | PPIB | 0.06 | Disease |

### 2.3 Interpreting protein and pathway relationships using the Reactome

To understand the main biological themes implicated by our results, we perform two additional analyses, focused on pathways and proteins, respectively. We consider how the specific selected pathways are related to each other and across analyses. To do so, we utilize the hierarchical structure of the Reactome database. Reactome establishes 29 “umbrella” pathways for Homo Sapiens, 21 of which are represented in the SomaScan data used for analysis. These umbrella pathways describe the main cellular processes (e.g., “Autophagy”), which are then subdivided into increasing granular cellular processes and reactions (see Fig. 2a for the first two levels of the Reactome hierarchy used in analysis.) The eight pathways that discriminated between the cases of CSF*α*synSAA+ and CSF*α*synSAA-fell into five umbrella pathways in Reactome: Immune System, Autophagy, Metabolism of Proteins, Extraceullar Matrix Organization, and Hemostasis (see Fig. 2b). Six umbrella pathways were associated with baseline mean striatum: Immune system, Metabolism of Proteins, Extracellular Matrix Organization, Signal Transduction, Autophagy, and Developmental Biology. Ten umbrella pathways were associated with trajectory: Immune System, Metabolism of Proteins, Extracellular Matrix Organization, Hemostasis, Cell-cell Communication, Disease, Cellular Responses to Stimuli, Signal Transduction, Transport of Small Molecules, and Metabolism. Thus, Immune System, Metabolism of Proteins, and Extracellular Matrix Organization were shared between all three analyses. Hemostasis, Signal Transduction, and Autophagy are shared between two analyses. Only the results for prediction of the trajectory of mean striatum had unique umbrella pathways, namely Cellular Response to Stimuli, Disease, Transport of Small Molecules, Sensory Perception and Gene Expression. Despite the fact that there is only one exact match between the selected pathways across the analysis (“VTN binds…” for CSF*α*synSAA and baseline mean striatum), by leveraging the hierarchical structure of Reactome, we are able to understand similarities of the discovered patterns. Motivated by the overlap in selected pathways across analyses, we select three, Immune System, Extracellular Matrix Organization, and Autophagy, to discuss in greater detail here.

**Figure 2.**
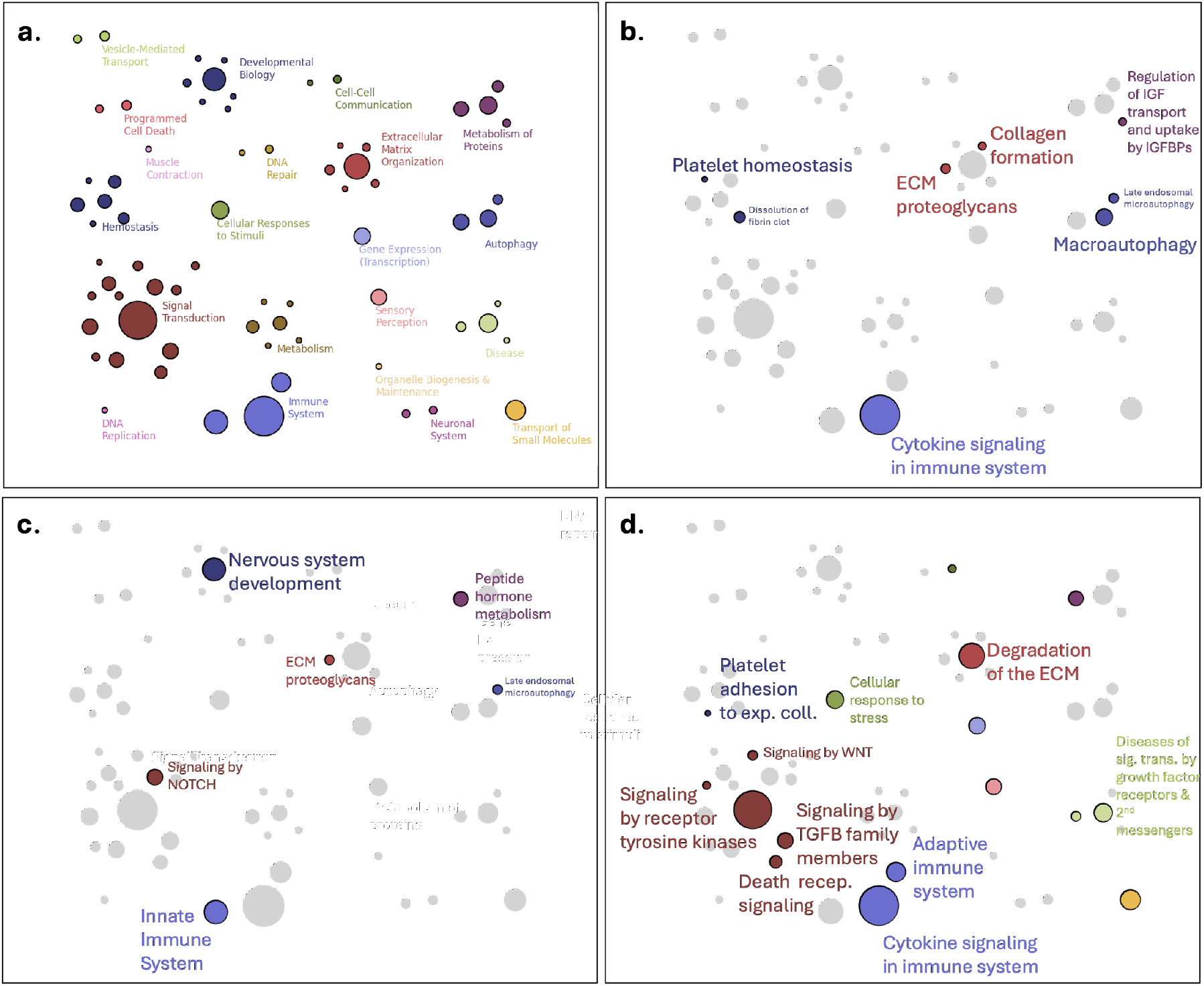
An overview of the candidate pathways and the selected pathways for the different analyses as organized by the first two-levels of Reactome annotations. **a.**Top-level organization of the 21 umbrella pathways in Reactome annotations which intersect the SomaScan measured proteins. The color represents the umbrella pathway, the number of circles per color represent the number of next level organization, and the size of the circle is proportional to the total number of pathways. **b**. The selected pathways for the CSF*α*synSAA status analysis. **c**. The selected pathways for the baseline mean striatum analysis. **d**. The selected pathways for the trajectory mean striatum analysis. For panels b-d, the top ten unique selections are annotated and the size of the font is proportional to the magnitude of the posterior inclusion probability. Immune System, Extracellular Matrix Organization, and Metabolism of Proteins are selected in all three analyses.

#### Immune System

The largest number of proteins and pathways we identified belonged to processes that are not part of the brain, but rather are part of the immune system. This includes many interleukins and interferons that have low levels of expression in brain [29]. There are multiple potential explanations for why the immune system may be important in *LRRK2*-associated parkinsonism. *LRRK2* is a ubiquitously expressed protein, with established roles in both peripheral immune function and autoimmunity [57, 54, 62, 43] as well as in microglia [53, 4, 45, 2]. Further, the finding of many immune-related proteins may reflect infiltration of peripheral immune cells into the brain, as occurs in PD [16, 61, 42] or expression in a subset of microglia [9]. Alternatively, these CSF proteins may have their origin in the periphery and can reflect a leaky blood brain barrier, active import from plasma to CSF, or failure of protein clearance from the CSF [19].

#### Extracellular Matrix Organization

We also identified several proteins and pathways involved in extracellular matrix organization. These include cathepsins (*CTSB, CTSL*), matrix metalloproteinases (*MMP3, MMP7*), integrins (*ITGB1, ITGB3, ITGB5*), and plasminogen (*PLG*). A third cathepsin, *CTSG*, was also identified in the Metabolism of Proteins pathway, and plasminogen was also identified in the Hemostasis pathway. Cathepsins are classically thought of as lysosomal proteases, and several studies have implicated *CTSB, CTSD*, and *CTSL* in PD pathogenesis [3, 26, 51, 27, 40]. Beyond their lysosomal function, cathepsins are also involved in tissue remodeling in the extracellular matrix [22], as well as in antigen presentation in the immune system [64]. Matrix metalloproteinases are extracellular matrix remodelers with many functions [30]. Plasminogen is canonically involved in the clotting cascade, but the plasminogen activation system is also expressed in the brain [24]. Integrins are receptors that allow cells to bind to extracellular proteins. They are especially critical for extravasation of lymphocytes from blood vessels. There are several ways in which extracellular matrix organization may be relevant to PD biology. Extracellular matrix organization is critical for microglia activation; thus we may speculate that these proteins are involved in neuroinflammation. Alternatively, it is possible that these genes are acting in neurons. In a recent paper in which the authors performed RNAseq on dopaminergic neurons that were differentiated from either healthy control or PD induced pluripotent stem cells (iPSC), extracellular matrix terms were among the top results that discriminated between the groups [50]. Finally, cathepsins, MMPs, and plasminogen have all been shown to cleave *α*-synuclein itself, so these processes may relate to control of *α*-synuclein, particularly in regards to extracellular spread [27, 34, 11, 52, 24].

#### Autophagy

Autophagy was identified in both the CSF*α*synSAA and baseline mean striatum analysis. Among the proteins contained in the selected pathways is *PARK7* (also known as *DJ-1*), which was increased in CSF*α*synSAA-cases relative to CSF*α*synSAA+. This is an exciting result, with clear relevance to PD, as loss of function mutations in *PARK7* are a known cause of autosomal recessive PD. *PARK7* functions as a chaperone protein and is sensitive to oxidative stress. A potential hypothesis arising from our findings would be that *LRRK2* cases that are CSF*α*synSAA-remain so because the increase in PARK7 offers them some protection from oxidative stress and allows them to better clear misfolded *α*-synuclein protein. In support of this hypothesis, in a Drosophila model of PD in which human *LRRK2* is over-expressed, additionally over-expressing human *PARK7* (*DJ-1*) reduced neurodegeneration in the fly eye [56]. Further, a more recent study using a toxic model of PD in the rat found that *PARK7* (*DJ-1*) and *LRRK2* independently regulate response to oxidative stress by acting through the same downstream pathway, which is activation of nuclear factor erythroid 2-related factor 2 (*Nrf2*) [44]. In this study, oxidative stress was worsened by *PARK7* (*DJ-1*) knockdown and rescued by *LRRK2* knockdown, consistent with our understanding of loss of function mutations of PARK7 causing PD versus gain of function *LRRK2* mutations causing PD. Collectively, these studies and our novel results lend support for the hypothesis that increasing *PARK7* (*DJ-1*) levels in *LRRK2*-associated parkinsonism would be helpful. This hypothesis should be tested further in future experiments.

### 2.4 Cell-type specific expression of nominated proteins

Protein abundance in CSF does not necessarily recapitulate protein expression within the brain parenchyma. Moreover, bulk proteomics provides only an averaged readout across a complex biological matrix, potentially obscuring signals arising from discrete or rare cellular populations. In contrast, single nuclei RNA sequencing (snRNA-seq) offers cell-type-resolved transcriptional profiles, enabling interrogation of molecular signatures within the cellular heterogeneity of diseased tissue. Although single-cell proteomics remains technically immature, available snRNA-seq datasets provide a useful orthogonal framework to ask whether proteins associated with CSF*α*synSAA status or mean striatum trajectory map to specific brain cell types—for example, whether inflammatory-associated markers preferentially localize to microglia.

To investigate this, we interrogated a publicly available snRNA-seq dataset from Kamanth et al., who profiled the substantia nigra from ten individuals with PD and eight controls [29]. This dataset captures transcriptional states across dopaminergic neurons, non-dopaminergic neurons, astrocytes, microglia, oligodendrocytes, oligodendrocyte progenitor cells, and endothelial cells (available at: https://singlecell.broadinstitute.org/single_cell/study/SCP1768/single-cell-genomic-profiling-of-human-dopamine-neurons-identifies-a-population-that-selectivelydegenerates-in-parkinsons-disease-single-nuclei-data).

Several notable patterns emerged. First, many proteins identified in our analyses were only minimally expressed across brain cell types in this dataset (see Figure 3), raising the possibility that a subset of CSF-associated signals may not originate from the brain parenchyma. Second, among genes with robust expression, most did not show strong cell-type restriction, suggesting that many candidate markers participate in broadly distributed rather than highly compartmentalized biological programs. Nonetheless, several exceptions were evident: *PDE2A, FLT3, PRDM4* and *FGF9* were enriched in neuronal populations; *AGT* was enriched in astrocytes; *FCGR2A* and *FGFR3* were preferentially expressed in microglia; and *ICAM2* was enriched in endothelial cells.

**Figure 3.**
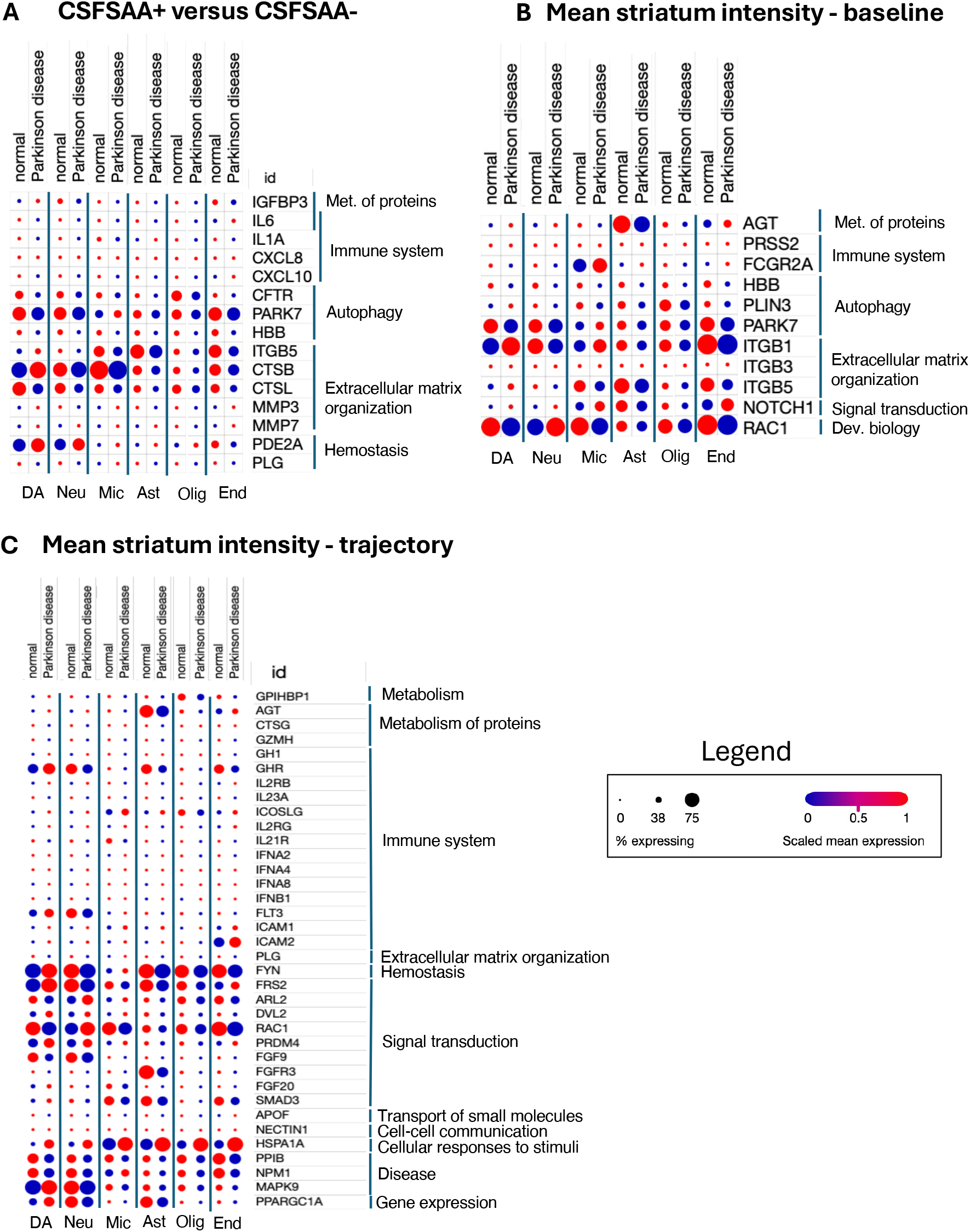
We used a publicly available single nucleus RNAseq study to examine cell-type specific expression of the genes identified in our analysis comparing CSF*α*synSAA+ and CSF*α*synSAA-cases (A) or those associated with mean striatium at baseline (B) or trajectory (C). The size of the circle is proportional to the percentage of each cell type that expressed the gene transcript. The color is proportional to the expression level of the gene transcript. Reactome umbrella pathways are indicated to the right of the individual genes. Many genes are not highly expressed in the brain in either PD or healthy controls. 17 genes demonstrate opposite changes in expression in PD in dopaminergic neurons compared to non-dopaminergic neurons.

Most strikingly, a substantial fraction of genes exhibited bidirectional, cell-type-dependent dysregulation in PD. *IGFBP3, ITGB5, CTSB, MMP3, PLIN3, ITGB1, GPIHBP1, GHR, FLT3, FYN, FRS2, FGF20, SMAD3, MAPK9, PPARGC1A* were increased in dopaminergic neurons but decreased in non-dopaminergic neurons, whereas *RAC1* and *ARL2* showed the opposite pattern. These opposing directional changes underscore the extent to which disease-associated molecular signatures are shaped by cellular context and suggest that aggregate CSF or bulk tissue measurements may collapse biologically divergent programs into deceptively uniform signals.

### 2.5 Comparison to conventional techniques

We compared the insights learned from the P-BANN approach to typical techniques in the field. L1-regularized, i.e. LASSO, models were fit for each of the phenotypes of interest (Methods Sec. 4.6). The linear model for CSF*α*synSAA had lower accuracy on the test set in both the protein and pathway settings, 67% and 73%, respectively. The models selected 77 proteins and 59 pathways. Using an enrichment analysis, we found no overlap between the selected proteins and pathways. In the case of the trajectory phenotype, using the proteins as input, the LASSO models selected 24 and 0 proteins for the intercept and slope, respectively. Using the pathways as input, the LASSO models selected 21 and 1 pathways for the intercept and slope, respectively. There is one statistically significant enriched pathway: DAPK binds UNC5B. This pathway is contained in the Programmed Cell Death hierarchy of the Reactome, and is a part of the Reactome that is never selected in the P-BANN analysis. Overall, the linear LASSO models have poorer predictive performance, no consistency between pathway- and protein-level analyses, and reveal a very limited amount of statistically significant features.

## 3 Discussion

P-BANN is a novel approach to discovering proteomic signatures guided-by phenotype. The design of P-BANN was guided by an interest in discovering a sparse set of proteins with connections that build on existing knowledge of protein-protein interactions via reactions and pathways. P-BANN can generally be applied to any setting with proteomics and an associated phenotype. Importantly, P-BANN can be used in combination with a variety of phenotype definitions, e.g. it is not restricted to binary outcomes. We demonstrated the utility of the technique via applications in PD.

The two defining features of PD are abnormal neuronal *α*-synuclein aggregates and loss of dopaminergic neurons in the substantia nigra [15]. The advent of biomarkers now allows their characterization *in vivo*, and *LRRK2* parkinsonism cases offer the opportunity to interrogate and contrast biologic pathways for each of these hallmark neuropathological features. Here we applied the P-BANN model to two separate questions: first to identify proteins and pathways that discriminate between *LRRK2* CSF*α*synSAA+ and CSF*α*synSAA-cases and second to identify proteins and pathways that are associated with progression of PD as measured by DAT SPECT imaging. Interestingly, while the individual pathways identified in each analysis were different, 3 umbrella pathways were present in all 3 analyses: Immune System, Metabolism of Proteins, and Extracellular Matrix Organization. Further, an additional 3 umbrella pathways (Autophagy, Signal Transduction, and Hemostasis) were shared between 2 out of 3 analyses. This suggests that there may be shared biological processes driving CSF*α*synSAA status and disease progression in *LRRK2*-associated parkinsonism.

P-BANN identified proteins of obvious relevance to PD (including *PARK7* and multiple cathepsins), but, importantly, this approach also allowed us to identify proteins in those same pathways that are of less obvious relevance, including *CFTR*, hemoglobin beta, matrix metalloproteinases, integrins, and plasminogen, many of which are targets of FDA approved drugs or are in drug development pipelines. Many of the proteins that we identified had a different direction of change in PD in dopaminergic neurons versus non-dopaminergic neurons, which could provide insights into the selective vulnerability of dopaminergic neurons. Collectively, these results shed light on the fundamental biology underlying PD and serve to generate hypotheses that can be confirmed in future experiments.

Our approach has some limitations. First, it is not possible at this time to validate our results in an independent cohort, as there is no other existing cohort of LRRK2-parkinsonism with biomarker data. Second, the applicability of our approach is limited by the availability of annotations and the coverage of proteomic assays. Both are growing, however, neither is comprehensive. In order to complete our study, we filtered the proteins measured by the SomaScan assay to include only those with coverage in the Reactome and which met our inclusion criteria (Methods) leading to a final dataset of 794 proteins. Using Reactome in this manner does inject some bias into the study. For instance, we expect that immune pathways are well studied and therefore well represented in the Reactome. There is likely inconsistent coverage, which could lead to missing or incorrect associations. However, we hypothesize that it is beneficial overall to include this information. This hypothesis is supported by the result that standard techniques did not result in the same biological insights and had poorer predictive performance overall. Importantly, our approach is not confined to any single annotation database, unlike past approaches [17].

We did not investigate alternative nonlinearities nor alternative distributional choices of phenotype as part of our study. These aspects of the P-BANN are modular and can be tuned to different applications of interest. This modularity is one of the particular benefits of the approach and specifically interrogating these modeling decisions is outside the scope of our study. We hope that others can build on this framework and share learnings concerning distributional and architectural design decisions.

In summary, we developed the P-BANN approach to be used in proteomic discovery. Its modular design, which allows for different annotation sources and different types of phenotypes, make it well suited to modern studies. We hope that the P-BANN approach will be a useful technique for a diverse set of future proteomic studies.

## 4 Methods

### 4.1 Study Sample

We selected the Parkinson’s Progression Markers Initiative (PPMI) cohort [33], a multi-center prospective study, for this analysis as it includes a large sample of individuals with *LRRK2* parkinsonism who have undergone CSF*α*synSAA testing, have CSF proteomics data available, and have longitudinal clinical and biomarker assessments. PPMI study methods have been described elsewhere in detail [33]. Briefly, as relevant to this analysis, PPMI recruited individuals diagnosed with PD based on clinical features, with disease duration of 7 years or less, who were known to have pathogenic variants in the *LRRK2* gene. Exclusion criteria were dementia and medical conditions that preclude study activities.

In this analysis we further excluded those who had (1) no CSF*α*synSAA result, (2) normal DAT SPECT binding in CSF*α*synSAA-individuals, (3) presence of known pathogenic GBA1 variant, and (4) an inconclusive or type II CSF*α*synSAA result.

Assessments include demographics, age at symptom onset, duration since clinical diagnosis at baseline visit, Movement Disorders Society Unified Parkinsons Disease Rating Scale (MDS-UPDRS), and levodopa equivalent daily dose (LEDD). Presence of aggregated asyn in CSF obtained at the baseline visit was assessed with the seed amplificaiton assay. Either of two versions of the assay may have been performed as described [8]. Test results were classified as positive (CSF*α*synSAA+) or negative (CSF*α*synSAA-) as described [13, 48]. Dopamine transporter (DAT) binding in the striatum was assessed with DATscan and SPECT as described [33]. SPECT scans were conducted at the baseline visit and at the year 2 and 4 assessments.

The CSF proteome was measured using the SomaScan aptamer-based proteomics platform. Data quality control and calibration were performed as described at ppmi-info.org (Project 151). The June 2024 release was used for analysis.

### 4.2 Modeling approach

We propose a modeling approach based on biologically-annotated neural networks (BANNs) [14]. BANN models were originally developed for association mapping in genomics data and aim to reflect multi-scale biology. This is achieved via a sparse neural network architecture design which aims to capture known biological relationships. BANNs are a variant of Bayesian neural networks and use spike-and-slab priors to enable feature selection. Here, we extend their definition to proteomics data, alter the parameterization of the prior, and consider non-binary definitions of phenotypes to guide association learning. In our setting, the architecture captures protein-protein interactions based on pathway annotations. This model definition is combined with a structured variational learning approach which enables generalization to classification, regression, and trajectory-based phenotypes.

Consider a setting with patients, each with 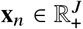 protein measurements and *y*_*n*_ ∈ ℝ ^*K*^ phenotype descriptor. We encode the relationship between the proteins and phenotype with an intermediate representation of *G* pathways as follows

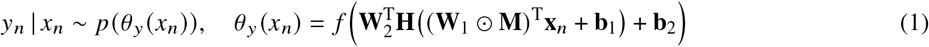

where 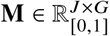 describes the mask which maps proteins to pathways, **H**(·) = [*h*_1_(·), …*h*_*G*_ (·)] is the set of non-linear activations applied to each pathway representation, and **W**_1_ ∈ ℝ^*J*×*G*^, **W**_2_ ∈ ℝ^*G*×*K*^, **b**_1_ ∈ ℝ^*G*^, and **b**_2_ ∈ ℝ^*K*^ are the weights and biases for the input and hidden layers, respectively. The functional forms of *p θ*_*y*_ *x*_*n*_ and *f* which maps the BANN to the distributional parameters depend on the phenotype of interest. Examples are given below.

As in standard BANNs, we aim to perform feature selection to enable proteomic signature discovery. To achieve this, we treat the weight parameters **W**_*l*_, with *l* ∈ {1, 2}, as random variables and apply sparsity-inducing prior distributions. We choose the horseshoe prior [6, 23], which is a continuous relaxation of the spike-and-slab prior. We describe each weight as follows using a Gaussian scale mixture

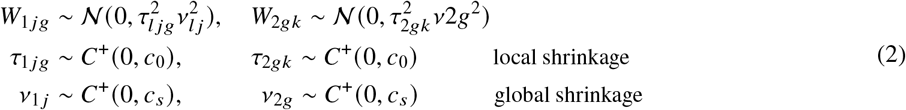

This choice of prior has been shown to be well-suited to feature selection tasks as sufficiently large weights remain un-shrunk, while small weights trend towards 0. This contrasts other choices of prior which have more uniform shrinkage [23].

The Half-Cauchy distribution present numerical challenges during variational learning but can be reparameterized using inverse-gamma distributions [58]. Using 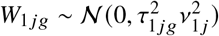 as an example,

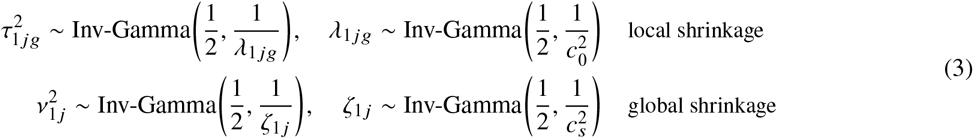

The parameterization of *W*_2*gk*_ follows analogously. This definition is further refined to use a non-center parameterization, also motivated by numerical properties [23]

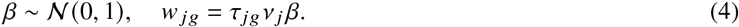

We complete the set of priors on weights by adding normal priors to the bias terms

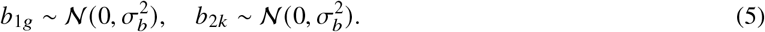

Defining all shrinkage parameters using 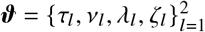, the joint distribution for our horseshoe BANN is

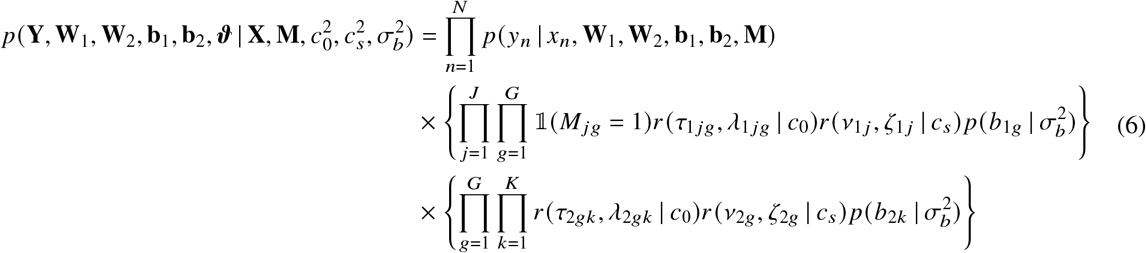

where *r* (*a, λ* | *c*) = Inv-Gamma (*a*^2^ | 1 / 2, 1 / *λ*) Inv-Gamma (*λ* | 1 / 2, 1 /*c* ^2^), 𝟙(*f* (*x*)) = 1 if (*f* (*x*) is True else 0, *N* is the number of samples, *J* is the number of proteins, *G* is the number of pathways, and *K* is the number of output nodes in the neural network.

Depending on the modeling question, we may wish to explicitly account for potential sources of confounding. Additional predictive covariates are straightforward to integrate into our approach. Instead of *θ*_*y*_ depending only on protein expression *x*_*n*_, we add additional measures *z* where

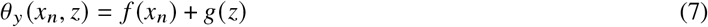

In this work, we consider linear functions for covariates where *g*(*z*) = *w*^T^*z* and we place normal priors over *w*.

As noted above, the definition of (*p y*_*n*_ | *x*_*n*_, **W**_1_, **W**_2_, **b**_1_, **b**_2_, **M**) depends on the phenotype. For instance, in the case of a binary phenotype, such that *y*_*n*_ ∈ {0, 1}, we use a Bernoulli random variable with probability *p* (*y*_*n*_ = 1 | *x*_*n*_) = *θ*_*y*_ (*x*_*n*_) and assume *f*(·) to be a sigmoid function. For the trajectory setting, *y*_*n*_ ∈ ℝ^*T*^ is modeled using a Gaussian process (GP) where *y*_*n*_ = *g*(*t*_*n*_, *x*_*n*_) + *ϵ* where *g* ∼ *GP* (*t, θ*_*y*_ (*x*_*n*_)) and *ϵ* ∼ 𝒩 (0, *σ*^2^). This modeling decision allows us to capture correlations in time, as opposed to modeling each time point independently, and increases data availability, as different patients have different number of visits (Table 4). By using black-box variational inference with unbiased Monte Carlo samples to estimate the intractable conditional distribution of *y*_*n*_, our framework is flexible to many definitions of phenotypic distribution. Further details on the learning approach and the specifics our the implementation for our research question can be found below.

**Table 4.** Demographic characteristics of the study sample at baseline.

|  | SAA+ Train<br>n=76 | SAA- Train<br>n=32 | SAA+ Test<br>n=19 | SAA- Test<br>n=9 |
| --- | --- | --- | --- | --- |
| Age at baseline (years) | 60.2 (8.4) | 68.3 (6.5) | 61.8 (8.8) | 69.9 (8.1) |
| Age at onset (years) | 54.9 (9.7) | 63.3 (6.5) | 57.2 (9.9) | 64.8 (9.9) |
| Disease duration (years) | 5.2 (6.5) | 5.0 (4.0) | 5.0 (3.0) | 5.2 (3.3) |
| Sex (male) | 43 (57%) | 10 (31%) | 12 (63%) | 5 (55%) |
| Race (% white) | 72 (95%) | 27 (84%) | 19 (100%) | 9 (100%) |
| *LRRK2* variant |  |  |  |  |
| N1437H | 1 (1%) | 0 (0%) | 0 (0%) | 0 (0%) |
| R1441G | 4 (5%) | 9 (28%) | 0 (0%) | 2 (22%) |
| R1441C | 1 (1%) | 0 (0%) | 0 (0%) | 0 (0%) |
| G2019S | 70 (92%) | 23 (72%) | 19 (100%) | 7 (78%) |
| I2020T | 0 (0%) | 0 (0%) | 0 (0%) | 0 (0%) |
| MDS-UPDRS Scores |  |  |  |  |
| MDS-UPDRS I | 7.9 (5.5) | 8.5 (6.9) | 7.7 (6.0) | 5.9 (3.8) |
| MDS-UPDRS II | 7.2 (4.9) | 7.0 (5.2) | 8.0 (4.1) | 4.1 (3.9) |
| MDS-UPDRS III - ON | 17.3 (9.5) | 18.6 (7.9) | 17.0 (9.1) | 15.6 (8.1) |
| Total MDS-UPDRS - ON | 32.4 (14.1) | 33.6 (16.0) | 33.6 (12.9) | 25.6 (11.2) |
| LEDD (mg) | 666 (468) | 304 (214) | 490 (220) | 388 (174) |
| Baseline Mean Striatum | 1.3 (0.4) | 1.3 (0.3) | 1.2 (0.4) | 1.5 (0.4) |
| Mean Striatum |  |  |  |  |
| 1 Visit | 26 (25%) |  | 8 (30%) |  |
| 2 Visit | 32 (30%) |  | 7 (26%) |  |
| 3 Visit | 43 (41%) |  | 10 (37%) |  |
| 4 Visit | 4 (4%) |  | 2 (7%) |  |

### 4.3 Learning approach

As noted above, we propose a structured variational approach to learning the posterior distribution. Specifically, as described in Ghosh et al. [23], we chose a fully-factorized variational distribution to approximate the posterior:

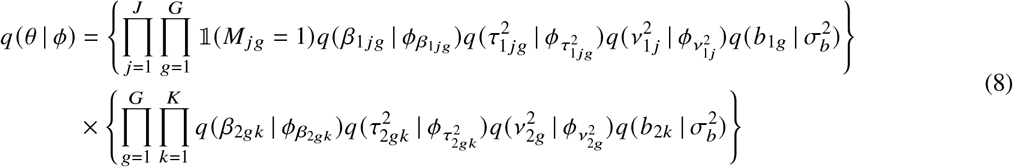

where (taking *l* ∈ {1, 2} as the layer index, and ignoring element indices) weight approximations are modeled using a Normal distribution 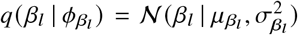, the bias is modeled using a Normal distribution 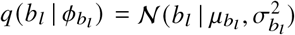, and both global and local shrinkage approximations are modeled using log-Normal distributions such that 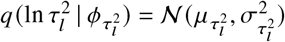 and 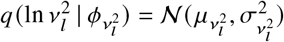.

Optimization proceeds by considering the evidence lower bound (ELBO) as the objective.

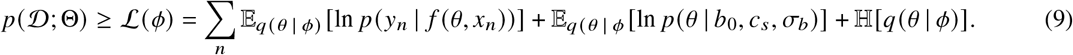

The intractable expectations are handled via unbiased sampling using the reparameterization trick.

### 4.4 Interrogating the posterior estimates

To understand the selection of proteins and pathways after learning, we leverage the posterior inclusion probability, which depends on the posterior shrinkage probability [6, 7]

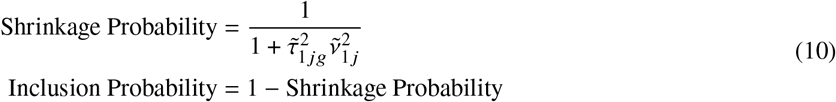

where 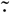· indicates the mean of the posterior and the definitions for the pathway layer are analogous.

### 4.5 Defining the model for Parkinson’s disease applications

Pathway annotations are selected based on the Reactome database [18] as exported on March 20, 2024. This yielded annotations for 11,662 proteins. An inclusion criterion was applied requiring a pathway to contain at least three participating proteins which decreased the sample to 3,711 proteins. Taking the intersection of this set with the proteins measured in the SomaScan panel yielded 1,789 proteins. Pathways were further refined to require at least three measured proteins and at least 50% of the proteins in the pathway to be measured. In the event two pathways had more than 50% overlap, the larger pathway was dropped from analysis.

A priori, we identified a set of proteins that are of high interest and priority based on their established relationship to PD (namely, proteins encoded by genes that are established to cause monogenic PD: *SNCA, PRKN, PINK1, PARK7, LRRK2, VPS35, GBA1*) and/or other neurodegenerative disorders (*GRN, C9orf72, MAPT, APOE, TARDBP*). In the case of these proteins, the overlap criteria is not applied. Importantly, this does not guarantee that these proteins or pathways are selected, only that they are not removed prior to completing the analysis. Although *C9orf27, PINK1, GBA*, and *VPS35* are proposed, they are not measured and are therefore excluded.

The final dataset contains 794 proteins and 407 pathways.

#### Specifying the phenotypic distributions

In this work, we present two analyses: (1) prediction of CSF*α*synSAA status (binary outcome) and (2) prediction of mean striatum trajectory up to five years. In the former, we model the phenotype as Bernoulli distribution such that

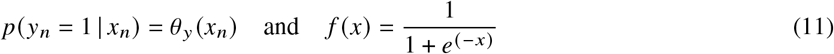

where *x* is a placeholder variable for the functional form described in eqn 1. In the latter, we model the phenotype using an amortized Gaussian process [38] such that

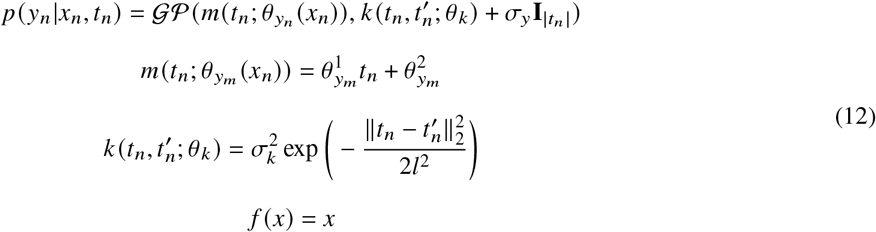

where each (*t*_*n*_, *y*_*n*_) trajectory is modeled as a Gaussian process with independent and identically distribution Gaussian noise. The selected mean function is linear and the mean and bias are the outputs of BANNs. The selected kernel function is squared exponential. The variance hyperparameters, *σ*_*y*_, *σ*_*k*_, and *l*, are fixed based on an initial analysis of the training data.

#### Additional covariates

Based on the prior literature [8], we select age and sex as additional covariates. Because of the potential correlations between age, sex and proteome, and the observed association between age, sex and SAA*α*synSAA status, we aimed to specifically allow the model to use these variables as covariates.

#### Optimization and prior selection

We summarize variables for the optimization and the selection of priors in Table 5.

**Table 5.**
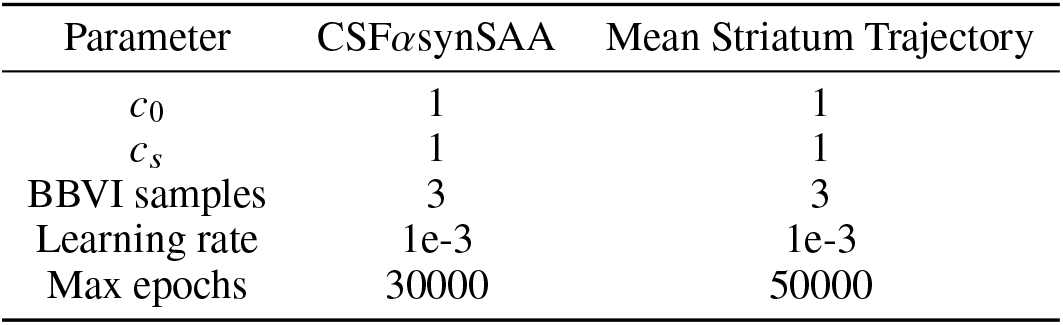
Prior values and optimization hyperparameters.

| Parameter | CSFαsynSAA | Mean Striatum Trajectory |
| --- | --- | --- |
| $c_0$ | 1 | 1 |
| $c_s$ | 1 | 1 |
| BBVI samples | 3 | 3 |
| Learning rate | 1e-3 | 1e-3 |
| Max epochs | 30000 | 50000 |

### 4.6 Baseline models

We consider two baseline approaches to compare to P-BANN. First, we perform a simple fold-change analysis for the CSF*α*synSAA+ and CSF*α*synSAA-protein expression with Bonferroni correction for multiple testing. Second, we consider L1-penalized models, also commonly called the LASSO [55]. We select this class of regularization technique because it is commonly used for feature selection, a key goal of our work.

For the classification of CSF*α*synSAA status, we perform L1-regularized logistic regression using the protein expression as input. In order to be able to compare with the pathway insights of the P-BANN, we also consider a setting where pathway annotations are used as a preprocessing step. Specifically, we calculate the average protein expression for a pathway and use this “pathway score” as an alternative input to the logistic regression.

There is not an obvious comparison technique for the trajectory phenotype. Therefore, as a baseline, we propose first fitting linear functions to each patient trajectory and then performed L1-regularized regression for each parameter, independently. This results in two sets of outcomes to interpret, similar to our P-BANN analysis, but different in that they are not learned jointly. As in the CSF*α*synSAA case, we use the protein expression as input as well as the mean protein expression per pathway.

In all settings, we consider the effect of including the covariates of age and sex, as is done in our P-BANN analysis as well as the effect of reweighted sampling for the classification task.

## 5 Data Availability

Data used in the preparation of this article were obtained from the Parkinson’s Progression Markers Initiative (PPMI) database (www.ppmi-info.org/access-data-specimens/download-data), RRID:SCR_006431.

## 6 Acknowledgments

This study was funded by Michael J Fox Foundation for Parkinson’s Research (MJFF).

A.V. is supported by UK Research and Innovation [UKRI Centre for Doctoral Training in AI for Healthcare grant number EP/S023283/1].

PPMI – a public-private partnership – is funded by the Michael J. Fox Foundation for Parkinson’s Research and funding partners, including 4D Pharma, Abbvie, AcureX, Allergan, Amathus Therapeutics, Aligning Science Across Parkinson’s, AskBio, Avid Radiopharmaceuticals, BIAL, BioArctic, Biogen, Biohaven, BioLegend, BlueRock Therapeutics, Bristol-Myers Squibb, Calico Labs, Capsida Biotherapeutics, Celgene, Cerevel Therapeutics, Coave Therapeutics, DaCapo Brainscience, Denali, Edmond J. Safra Foundation, Eli Lilly, Gain Therapeutics, GE HealthCare, Genentech, GSK, Golub Capital, Handl Therapeutics, Insitro, Jazz Pharmaceuticals, Johnson & Johnson Innovative Medicine, Lundbeck, Merck, Meso Scale Discovery, Mission Therapeutics, Neurocrine Biosciences, Neuron23, Neuropore, Pfizer, Piramal, Prevail Therapeutics, Roche, Sanofi, Servier, Sun Pharma Advanced Research Company, Takeda, Teva, UCB, Vanqua Bio, Verily, Voyager Therapeutics, the Weston Family Foundation and Yumanity Therapeutics.

